# A model-based approach to identifying atypical type 2 diabetes: UK population-based study

**DOI:** 10.64898/2026.11.20.26368556

**Authors:** MM Dinsdale, AP McGovern, P Cardoso, LM Güdemann, TT Jansz, AG Jones, AT Hattersley, TJ McKinley, BM Shields, KG Young, JM Dennis

**Author notes:** **Corresponding author:** John M. Dennis, Address: Clinical and Biomedical Sciences, University of Exeter Medical School, Exeter, United Kingdom.

## Abstract

**Aims:** Approaches to meaningfully identify individuals with differing prognosis at type 2 diabetes diagnosis are lacking. We aimed to assess whether QDiabetes, a validated routine clinical feature-based model to predict risk of type 2 diabetes onset, could be applied at the point of diagnosis to identify people with atypical type 2 diabetes with differing risk of complications and treatment requirements.

**Methods:** We included adults newly diagnosed with type 2 diabetes between 2013–2023 in population-representative UK primary care data (Clinical Practice Research Datalink). Individuals with low predicted QDiabetes risk (≤5.6%) were classified as atypical type 2 diabetes and matched up to 1:4 by age, sex and calendar year to typical type 2 diabetes (QDiabetes risk >5.6%). We compared 10-year risks of microvascular, macrovascular, acute complications, mortality and glucose-lowering treatment initiation. For complications, multivariable Cox models estimated adjusted hazard ratios (aHRs), adjusting for age, sex, year of diagnosis, ethnicity, deprivation and baseline HbA1c.

**Results:** Of 276,117 individuals newly diagnosed with type 2 diabetes, 30,397 (11.0%) were classified as atypical type 2 diabetes using our approach. Compared with the wider typical type 2 diabetes population (n=245,720; 89.0%), they were younger (54 vs 60 years), leaner (26.4 vs 34.0 kg/m²) and had higher HbA1c at diagnosis (68.0 vs 60.0 mmol/mol). In matched analyses, the atypical type 2 diabetes group had a higher risk of severe retinopathy (aHR 1.33, 95% CI 1.23–1.44), similar risk of severe neuropathy (aHR 0.92, 0.84–1.01), and lower risk of severe nephropathy (aHR 0.81, 0.73–0.90). Despite lower predicted baseline cardiovascular risk in those with atypical type 2 diabetes, risks of myocardial infarction (aHR 0.99, 0.89–1.11) and stroke (aHR 0.92, 0.81–1.04) were similar, while heart failure risk was lower (aHR 0.72, 0.67–0.78). Atypical type 2 diabetes was also associated with higher risks of diabetic ketoacidosis, severe hypoglycaemia, non-cardiovascular mortality and insulin initiation.

**Conclusion:** Type 2 diabetes onset risk prediction models used at the point of diagnosis may represent a pragmatic approach to identify people with an atypical type 2 diabetes phenotype. Despite fewer conventional risk factors, the phenotype identified had a poor prognosis and could provide a practical target for personalised type 2 diabetes management to reduce risk of long-term complications.

## INTRODUCTION

Type 2 diabetes is a heterogeneous condition with marked variation in disease progression.^1–3^ Phenotypic variation is apparent at clinical diagnosis and reflects a complex interplay of genetic, environmental and lifestyle factors.^4^ A better understanding of heterogeneity in type 2 diabetes at diagnosis could support more individualised management and improve long-term outcomes.^1 5 6^ Although recent approaches have proposed subclassification of type 2 diabetes into clinically meaningful phenotypes, these have yet to translate into actionable groups in routine practice.^7 8^

Risk prediction models to predict type 2 diabetes onset using routinely available features may offer a pragmatic way to identify individuals at diagnosis with differences in subsequent risk of complications. Risk prediction models including QDiabetes, FINDRISC and the American Diabetes Association Diabetes Risk Test are increasingly used to predict individuals at high risk of developing type 2 diabetes and to guide targeted glycaemic diagnostic testing.^9–11^ They estimate risk from established predictors such as age, body mass index, ethnicity, family history and comorbidities. Individuals who develop type 2 diabetes despite a low predicted risk are therefore likely to have fewer conventional risk factors and may represent an atypical phenotype. However, the clinical characteristics, disease progression, and treatment requirements of such an atypical group have not previously been explored.

We aimed to assess the characteristics, prognosis, and treatment of individuals who developed type 2 diabetes despite low predicted risk of developing diabetes (atypical type 2 diabetes phenotype), compared with age- and sex-matched individuals with high predicted risk (typical type 2 diabetes phenotype). Following diagnosis, we examined the risks of severe microvascular and macrovascular complications, and initiation of glucose-lowering treatment.

## METHODS

### Study population

We used longitudinal UK primary care data from Clinical Practice Research Datalink (CPRD) Aurum.^12^ Primary care records were linked by CPRD to secondary care records (Hospital Episode Statistics [HES]), deprivation records (2019 English Index of Multiple Deprivation^13^), and death certificate records (Office for National Statistics). CPRD is population representative and covers approximately 20% of the UK population.^14^

We included adults with valid HES linkage aged 25 to 84 years with newly diagnosed type 2 diabetes between January 2013 and March 2023, requiring both a recorded type 2 diabetes diagnostic code and a confirmatory HbA1c ≥48 mmol/mol. The index date for study follow-up was the assigned date of type 2 diabetes diagnosis, defined as the earliest diagnostic code, HbA1c ≥48 mmol/mol, or initiation of glucose-lowering treatment in the primary care record. We excluded individuals if they had evidence of type 1 diabetes (insulin initiation within 1 year of diagnosis or any recorded type 1 diabetes code), type 3c diabetes (history of acute/chronic pancreatitis, pancreatic cancer, pancreatic resection or haemochromatosis), or cystic fibrosis.

### Covariates

Baseline characteristics and comorbidities were defined at or before the date of type 2 diabetes diagnosis using primary care (Read, SNOMED-CT) and hospital inpatient (ICD-10, OPCS-4) codes (codelists available at https://github.com/Exeter-Diabetes/CPRD-Codelists). Demographic variables included age at diagnosis, sex, ethnicity (categorised according to major UK census categories: White, South Asian, Black, Mixed, Other), social deprivation (IMD quintile with 1 being least deprived and 5 most deprived), smoking status, alcohol use and family history of diabetes.

Clinical biomarkers included HbA1c (mmol/mol), BMI (kg/m²), eGFR (mL/min/1.73 m²), HDL cholesterol (mmol/L), LDL cholesterol (mmol/L) and triglyceride-to-HDL cholesterol ratio (used as a routinely available surrogate marker of insulin resistance).^15^ Biomarker values were taken closest to diagnosis within the previous 2 years, except for HbA1c (within the previous 6 months) and BMI (from 5 years before to 1 year after diagnosis to maximise completeness).

Baseline comorbidities, informed by existing diabetes risk scores,^9 11 16 17^ included cardiovascular disease (myocardial infarction, stroke, revascularisation, ischaemic heart disease, angina, peripheral arterial disease, or transient ischaemic attack), heart failure, hypertension requiring treatment, chronic kidney disease stages 3–5, polycystic ovary syndrome, bipolar disorder or schizophrenia, learning disabilities and gestational diabetes. In those without pre-existing cardiovascular disease (CVD), baseline 10-year cardiovascular risk was estimated using the QRISK2-2008 10-year score.^16^ Medication history at diagnosis included statins, oral steroids, and antipsychotics.

### Definition of atypical type 2 diabetes

To define an atypical type 2 diabetes group, the QDiabetes-2018a risk algorithm was applied at the date of diabetes diagnosis.^9^ QDiabetes was originally developed to predict 10-year risk of developing type 2 diabetes and therefore it would be anticipated that those who developed diabetes despite low predicted risk would represent an atypical type 2 diabetes phenotype.

QDiabetes-2018a inputs comprised age, sex, ethnicity, Townsend deprivation, BMI, smoking status, family history of diabetes, cardiovascular disease, treated hypertension, corticosteroid use, atypical antipsychotic use, statin use, schizophrenia or bipolar disorder, learning disability, gestational diabetes and polycystic ovary syndrome. HbA1c is not included in QDiabetes-2018a as the primary purpose of the model is to identify high-risk individuals suitable for HbA1c testing. A valid QDiabetes score was calculated only when all required predictor variables were complete and within permitted ranges; individuals without a valid score were excluded (n=43,305, 13.6%; Supplementary Flowchart). Individuals with a QDiabetes score ≤5.6% were classified as having an atypical phenotype, corresponding to below the UK NHS Health Check threshold for high diabetes risk (>5.6%).^18^

### Study outcomes

#### Primary outcomes

Primary outcomes were severe microvascular and macrovascular complications following type 2 diabetes diagnosis, identified from primary care, secondary care and mortality records.

Microvascular complications included severe retinopathy (vitreous or pre-retinal haemorrhage, photocoagulation, proliferative retinopathy, blindness or visual impairment), severe neuropathy (foot ulcer, gastroparesis, Charcot foot, painful peripheral neuropathy, neuropathic pain or lower-limb amputation), and severe nephropathy (sustained ≥40% decline in eGFR, end-stage kidney disease [renal replacement therapy or sustained eGFR <15 mL/min/1.73 m²], or death from kidney failure).

Macrovascular complications comprised fatal or non-fatal myocardial infarction (MI), stroke and heart failure.

#### Secondary outcomes

Secondary outcomes included acute complications comprising diabetic ketoacidosis and severe hypoglycaemia (identified from primary cause of hospital admission records), mortality (cardiovascular and non-cardiovascular), and post-diagnosis treatment initiation.

Treatment outcomes comprised initiation of insulin, first-line glucose-lowering therapy, and second-line therapy added to initial metformin. Glucose-lowering therapies were identified from primary care prescribing records and included metformin, sulphonylureas, DPP-4 inhibitors, SGLT2 inhibitors, GLP-1 receptor agonists, thiazolidinediones, glinides and acarbose.

### Statistical analysis

We aimed to compare the risk of study outcomes between individuals with atypical and typical type 2 diabetes. Each person in the atypical type 2 diabetes group was matched to up to four people with a high predicted diabetes risk score (QDiabetes risk score >5.6%), exactly matched on age at diabetes diagnosis (rounded to the nearest year), sex and calendar year of diagnosis. Matching was performed with replacement. Standardised mean differences (SMDs) were used to assess balance in baseline characteristics after matching.

#### Risk of new onset diabetes complications, mortality and treatment

We estimated 10-year cumulative incidence for each complication outcome using Kaplan-Meier curves. Follow-up began at type 2 diabetes diagnosis and continued until the earliest of: complication occurrence, date of practice deregistration/death, end of study period (March 2023, which was the end of available follow-up in HES). For each outcome analysis, individuals with a history of the outcome of interest at diagnosis were excluded, as well as their matched comparators.

Cox proportional hazards models estimated hazard ratios comparing those with atypical type 2 diabetes versus their matched counterparts. The primary analysis adjusted for demographic confounders including age at diabetes diagnosis, sex, calendar year of diagnosis, ethnicity and deprivation, and baseline HbA1c to additionally adjust for the mediating effect of HbA1c at diagnosis. This analysis therefore assessed if differences between the groups were independent of glycaemia at diagnosis, which may relate to both timing of diagnosis as well as underlying clinical phenotype. Missing baseline HbA1c was imputed using multiple imputation by chained equations with 10 imputed datasets. Estimates were pooled according to Rubin’s rules. Proportional hazards assumptions were assessed visually using Schoenfeld residuals.^19^ Consistency of associations were finally assessed across subgroups defined by age at diagnosis (<50 or ≥50 years) and sex.

For treatment outcomes, we estimated 10-year cumulative incidence of treatment initiations, with relative effects estimated using unadjusted hazard ratios. We did not adjust for baseline HbA1c as this is likely to be the primary determinant of an individual initiating therapy. Follow-up for insulin initiation began 1 year after diagnosis.

All statistical analyses used R (v4.4.0).^20^ We followed RECORD reporting guidelines.^21^

## RESULTS

### Clinical characteristics of model-defined atypical type 2 diabetes group

Among 276,117 study-eligible individuals newly diagnosed with type 2 diabetes between 2013 and 2023, 30,397 (11.0%) developed diabetes despite a low predicted diabetes risk (QDiabetes score ≤5.6%) and were classified as having atypical type 2 diabetes (Supplementary Flowchart). Compared with the typical type 2 diabetes population with a higher QDiabetes score at diagnosis (QDiabetes score >5.6%; n=245,720), individuals with atypical type 2 diabetes were younger at diagnosis (mean age 54 vs 60 years), leaner (mean BMI 26.4 vs 34.0 kg/m²), and had higher HbA1c (68.0 vs 60.0 mmol/mol) (Supplementary Figure 1). However, atypical type 2 diabetes occurred across a broad range of age and BMI: 54.9% were aged ≥50 years and 15.8% had a BMI ≥30 kg/m² (Supplementary Table 1). Mean study follow-up was 4.4 (SD 2.9) years in the atypical type 2 diabetes and 4.5 (SD 2.9) years in the typical type 2 diabetes population.

Atypical type 2 diabetes had a similar proportion of White individuals (79.9% vs 78.7%) but a slightly lower proportion of South Asian individuals (9.5% vs 12.9%) than the majority typical type 2 diabetes population. Atypical type 2 diabetes was also relatively evenly distributed across deprivation quintiles (21.6% in the least deprived vs 18.0% in the most deprived), whereas a higher proportion of typical type 2 diabetes was in the most deprived than the least deprived quintile (25.1% vs 15.3%).

Individuals with atypical type 2 diabetes had a more favourable baseline cardiovascular risk profile, with a lower prevalence of cardiovascular disease (7.4% vs 22.2%) and, in those without cardiovascular disease, lower mean QRISK2 scores (10-year cardiovascular disease risk 16.7% vs 25.4%). They were less likely to be active smokers (20.9% vs 23.2%) and to receive statins (11.0% vs 39.1%). Mean triglyceride-to-HDL cholesterol ratio was similar between groups (2.3 vs 2.3), as was recorded family history of diabetes (15.6% vs 15.1%).

After matching on age, sex and calendar year of diagnosis, 30,392 individuals with atypical type 2 diabetes were matched to 121,294 individuals with typical type 2 diabetes. While age and sex were balanced after matching (SMD <0.01), the broader pattern of clinical differences persisted, including lower BMI, higher HbA1c and a more favourable cardiovascular risk profile, including lower QRISK2 scores (16.7% vs 20.1%), lower cardiovascular disease prevalence (7.4% vs 16.9%) and less treated hypertension (11.1% vs 38.0%) (Table 1). Mean follow-up was 4.5 (SD 2.9) years in the matched typical type 2 diabetes group.

**Table 1.**
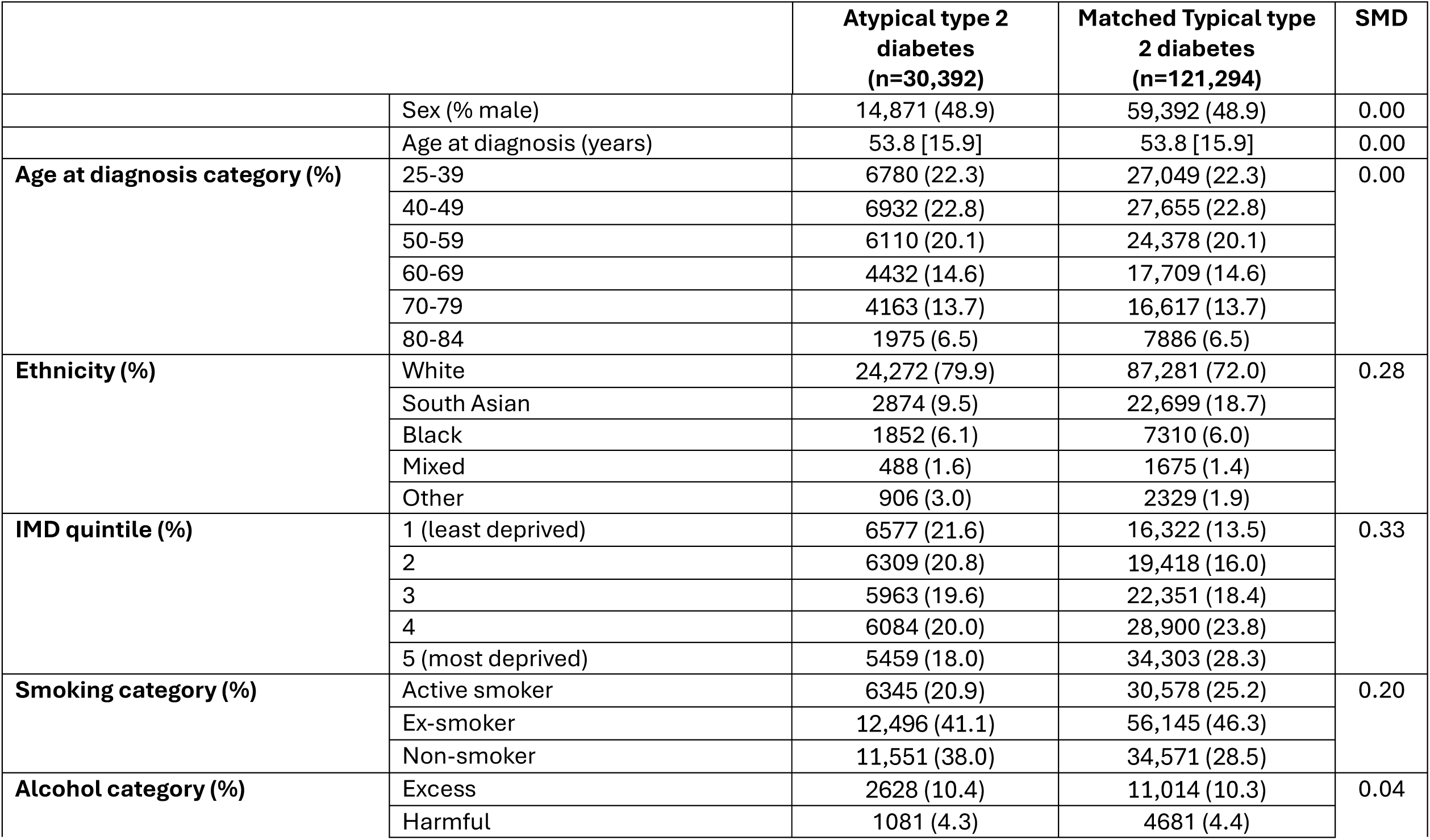
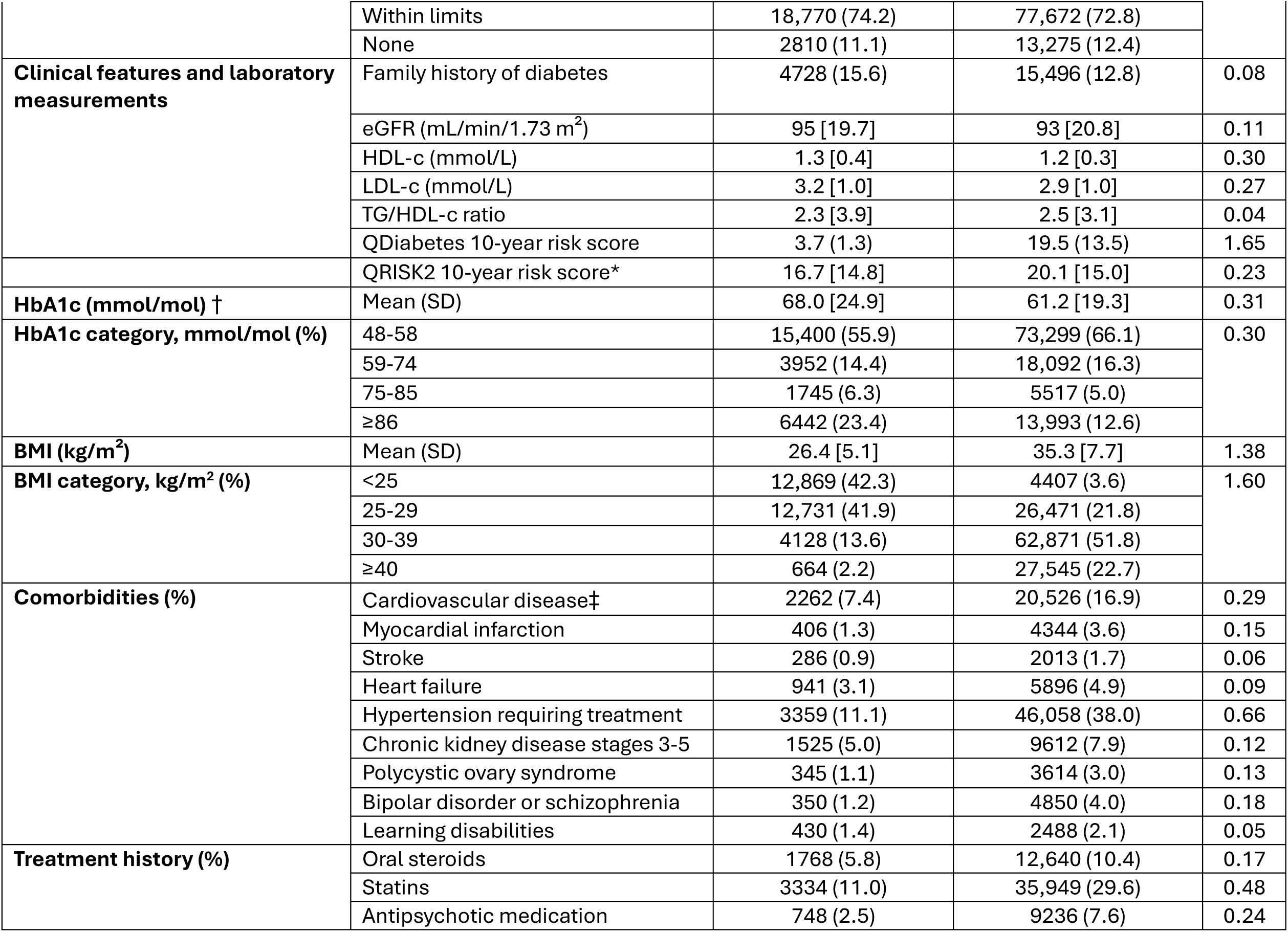
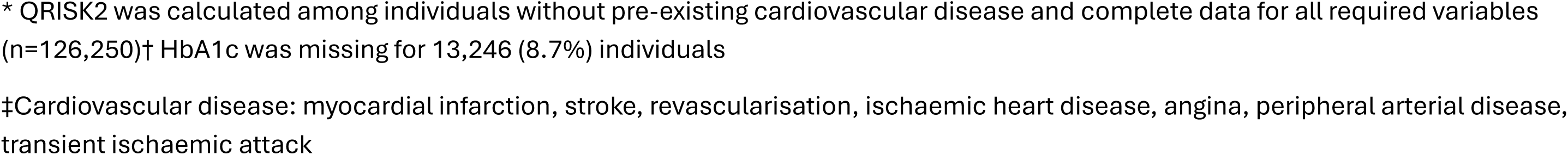
Baseline characteristics in atypical versus matched typical type 2 diabetes. Individuals with atypical type 2 diabetes (QDiabetes risk score ≤5.6%; n=30,392) were compared with age-, sex- and calendar year-matched individuals with typical type 2 diabetes (QDiabetes risk score >5.6%; n=121,294). Values are n (%) for categorical variables and mean [SD] for continuous variables. Percentages for variables with missing data are calculated among individuals with available data. Standardised mean differences (SMDs) compare the atypical type 2 diabetes group with the matched typical type 2 diabetes group, SMD >0.1 indicates meaningful imbalance.

|  |  | Atypical type 2 diabetes<br>(n=30,392) | Matched Typical type 2 diabetes<br>(n=121,294) | SMD |
|---|---|---|---|---|
|  | Sex (% male) | 14,871 (48.9) | 59,392 (48.9) | 0.00 |
|  | Age at diagnosis (years) | 53.8 [15.9] | 53.8 [15.9] | 0.00 |
| **Age at diagnosis category (%)** | 25-39 | 6780 (22.3) | 27,049 (22.3) | 0.00 |
|  | 40-49 | 6932 (22.8) | 27,655 (22.8) |  |
|  | 50-59 | 6110 (20.1) | 24,378 (20.1) |  |
|  | 60-69 | 4432 (14.6) | 17,709 (14.6) |  |
|  | 70-79 | 4163 (13.7) | 16,617 (13.7) |  |
|  | 80-84 | 1975 (6.5) | 7886 (6.5) |  |
| **Ethnicity (%)** | White | 24,272 (79.9) | 87,281 (72.0) | 0.28 |
|  | South Asian | 2874 (9.5) | 22,699 (18.7) |  |
|  | Black | 1852 (6.1) | 7310 (6.0) |  |
|  | Mixed | 488 (1.6) | 1675 (1.4) |  |
|  | Other | 906 (3.0) | 2329 (1.9) |  |
| **IMD quintile (%)** | 1 (least deprived) | 6577 (21.6) | 16,322 (13.5) | 0.33 |
|  | 2 | 6309 (20.8) | 19,418 (16.0) |  |
|  | 3 | 5963 (19.6) | 22,351 (18.4) |  |
|  | 4 | 6084 (20.0) | 28,900 (23.8) |  |
|  | 5 (most deprived) | 5459 (18.0) | 34,303 (28.3) |  |
| **Smoking category (%)** | Active smoker | 6345 (20.9) | 30,578 (25.2) | 0.20 |
|  | Ex-smoker | 12,496 (41.1) | 56,145 (46.3) |  |
|  | Non-smoker | 11,551 (38.0) | 34,571 (28.5) |  |
| **Alcohol category (%)** | Excess | 2628 (10.4) | 11,014 (10.3) | 0.04 |
|  | Harmful | 1081 (4.3) | 4681 (4.4) |  |
|  | Within limits | 18,770 (74.2) | 77,672 (72.8) |  |
|  | None | 2810 (11.1) | 13,275 (12.4) |  |
| **Clinical features and laboratory measurements** | Family history of diabetes | 4728 (15.6) | 15,496 (12.8) | 0.08 |
|  | eGFR (mL/min/1.73 m<sup>2</sup>) | 95 [19.7] | 93 [20.8] | 0.11 |
|  | HDL-c (mmol/L) | 1.3 [0.4] | 1.2 [0.3] | 0.30 |
|  | LDL-c (mmol/L) | 3.2 [1.0] | 2.9 [1.0] | 0.27 |
|  | TG/HDL-c ratio | 2.3 [3.9] | 2.5 [3.1] | 0.04 |
|  | QDiabetes 10-year risk score | 3.7 (1.3) | 19.5 (13.5) | 1.65 |
|  | QRISK2 10-year risk score* | 16.7 [14.8] | 20.1 [15.0] | 0.23 |
| **HbA1c (mmol/mol) †** | Mean (SD) | 68.0 [24.9] | 61.2 [19.3] | 0.31 |
| **HbA1c category, mmol/mol (%)** | 48-58 | 15,400 (55.9) | 73,299 (66.1) | 0.30 |
|  | 59-74 | 3952 (14.4) | 18,092 (16.3) |  |
|  | 75-85 | 1745 (6.3) | 5517 (5.0) |  |
|  | ≥86 | 6442 (23.4) | 13,993 (12.6) |  |
| **BMI (kg/m<sup>2</sup>)** | Mean (SD) | 26.4 [5.1] | 35.3 [7.7] | 1.38 |
| **BMI category, kg/m<sup>2</sup> (%)** | <25 | 12,869 (42.3) | 4407 (3.6) | 1.60 |
|  | 25-29 | 12,731 (41.9) | 26,471 (21.8) |  |
|  | 30-39 | 4128 (13.6) | 62,871 (51.8) |  |
|  | ≥40 | 664 (2.2) | 27,545 (22.7) |  |
| **Comorbidities (%)** | Cardiovascular disease‡ | 2262 (7.4) | 20,526 (16.9) | 0.29 |
|  | Myocardial infarction | 406 (1.3) | 4344 (3.6) | 0.15 |
|  | Stroke | 286 (0.9) | 2013 (1.7) | 0.06 |
|  | Heart failure | 941 (3.1) | 5896 (4.9) | 0.09 |
|  | Hypertension requiring treatment | 3359 (11.1) | 46,058 (38.0) | 0.66 |
|  | Chronic kidney disease stages 3-5 | 1525 (5.0) | 9612 (7.9) | 0.12 |
|  | Polycystic ovary syndrome | 345 (1.1) | 3614 (3.0) | 0.13 |
|  | Bipolar disorder or schizophrenia | 350 (1.2) | 4850 (4.0) | 0.18 |
|  | Learning disabilities | 430 (1.4) | 2488 (2.1) | 0.05 |
| **Treatment history (%)** | Oral steroids | 1768 (5.8) | 12,640 (10.4) | 0.17 |
|  | Statins | 3334 (11.0) | 35,949 (29.6) | 0.48 |
|  | Antipsychotic medication | 748 (2.5) | 9236 (7.6) | 0.24 |

\* QRISK2 was calculated among individuals without pre-existing cardiovascular disease and complete data for all required variables (n=126,250)† HbA1c was missing for 13,246 (8.7%) individuals
‡Cardiovascular disease: myocardial infarction, stroke, revascularisation, ischaemic heart disease, angina, peripheral arterial disease, transient ischaemic attack

### Model-defined atypical type 2 diabetes group has a higher incidence of some adverse outcomes including severe retinopathy

For microvascular complications, the 10-year incidence of severe retinopathy was higher in atypical than matched typical type 2 diabetes (8.2% [95% CI 7.5 to 9.0] vs 6.1% [5.8 to 6.5]) (Figure 1). Severe neuropathy incidence was similar between groups (6.0% [5.4 to 6.7] vs 6.9% [6.6 to 7.3]), while severe nephropathy incidence was lower in atypical type 2 diabetes (5.0% [4.4 to 5.7] vs 6.5% [6.1 to 6.8]).

**Figure 1.**
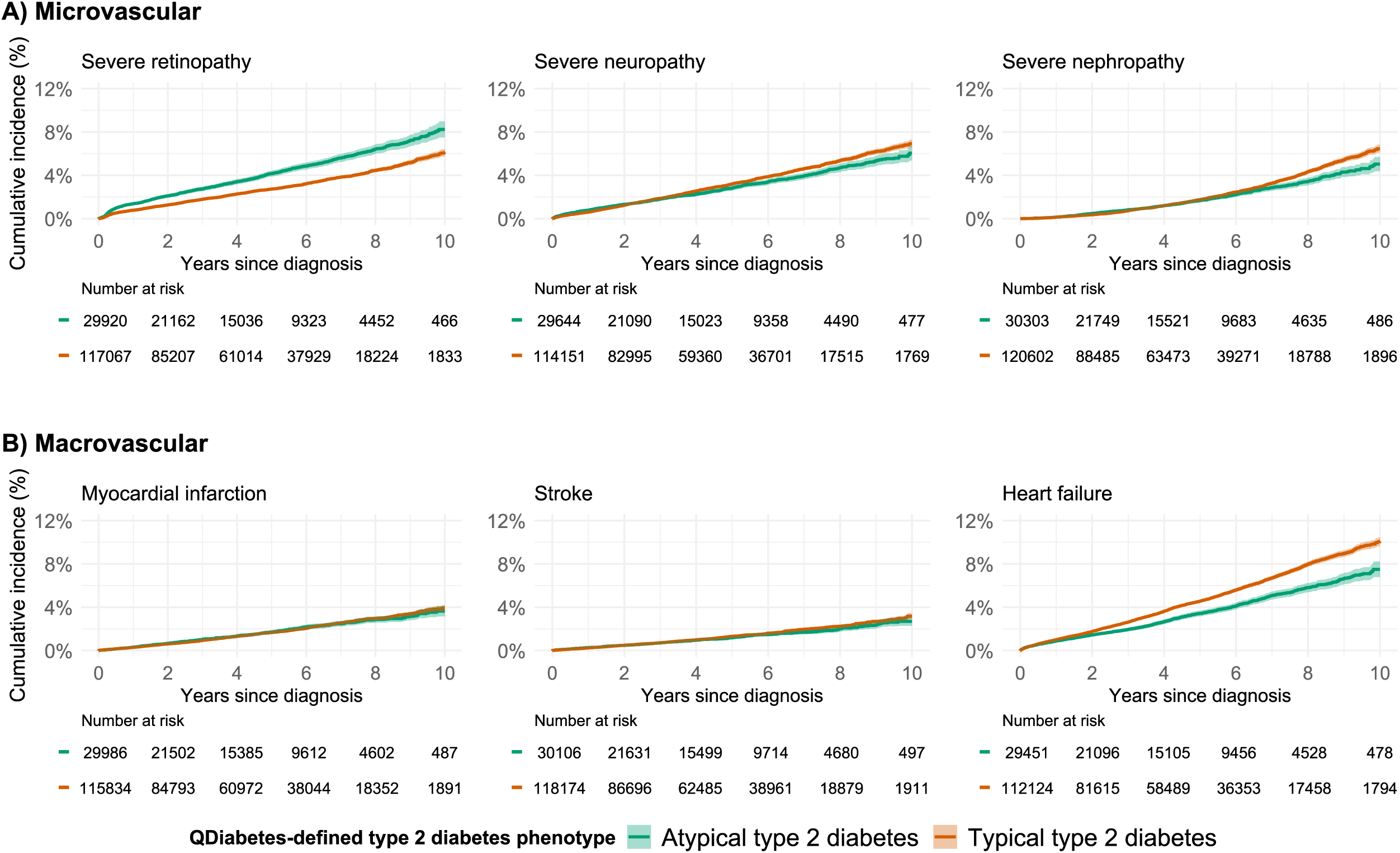
Ten-year cumulative incidence of primary complications in atypical versus matched typical type 2 diabetes. Kaplan–Meier curves compare individuals with atypical type 2 diabetes (QDiabetes risk score ≤5.6%; n=30,392) with age-, sex- and calendar year-matched individuals with typical type 2 diabetes (QDiabetes risk score >5.6%; n=121,294). Outcomes were microvascular complications (severe retinopathy, neuropathy and nephropathy) and macrovascular complications (fatal or non-fatal myocardial infarction, stroke and heart failure). Shaded areas indicate 95% confidence intervals; numbers below each panel indicate participants at risk.

For macrovascular complications, despite lower baseline QRISK2 score, 10-year incidence was similar between atypical and matched typical type 2 diabetes for myocardial infarction (3.6% [3.1 to 4.1] vs 3.9% [3.7 to 4.2]) and stroke (2.7% [2.3 to 3.1] vs 3.2% [2.9 to 3.4]). Heart failure incidence was lower in atypical type 2 diabetes (7.5% [6.8 to 8.2] vs 10.1% [9.7 to 10.5]).

### Differences in complication risk persist after adjustment for sociodemographic factors and mediating effect of baseline HbA1c

After adjustment for age at diagnosis, sex, calendar year of diagnosis, ethnicity, deprivation and baseline HbA1c, atypical type 2 diabetes remained associated with a higher risk of severe retinopathy (aHR 1.33, 95% CI 1.23 to 1.44) (Figure 2; Supplementary Table 2). Severe neuropathy risk was similar between groups (aHR 0.92, 95% CI 0.84 to 1.01), while severe nephropathy risk was lower in atypical type 2 diabetes (aHR 0.81, 95% CI 0.73 to 0.90).

**Figure 2.**
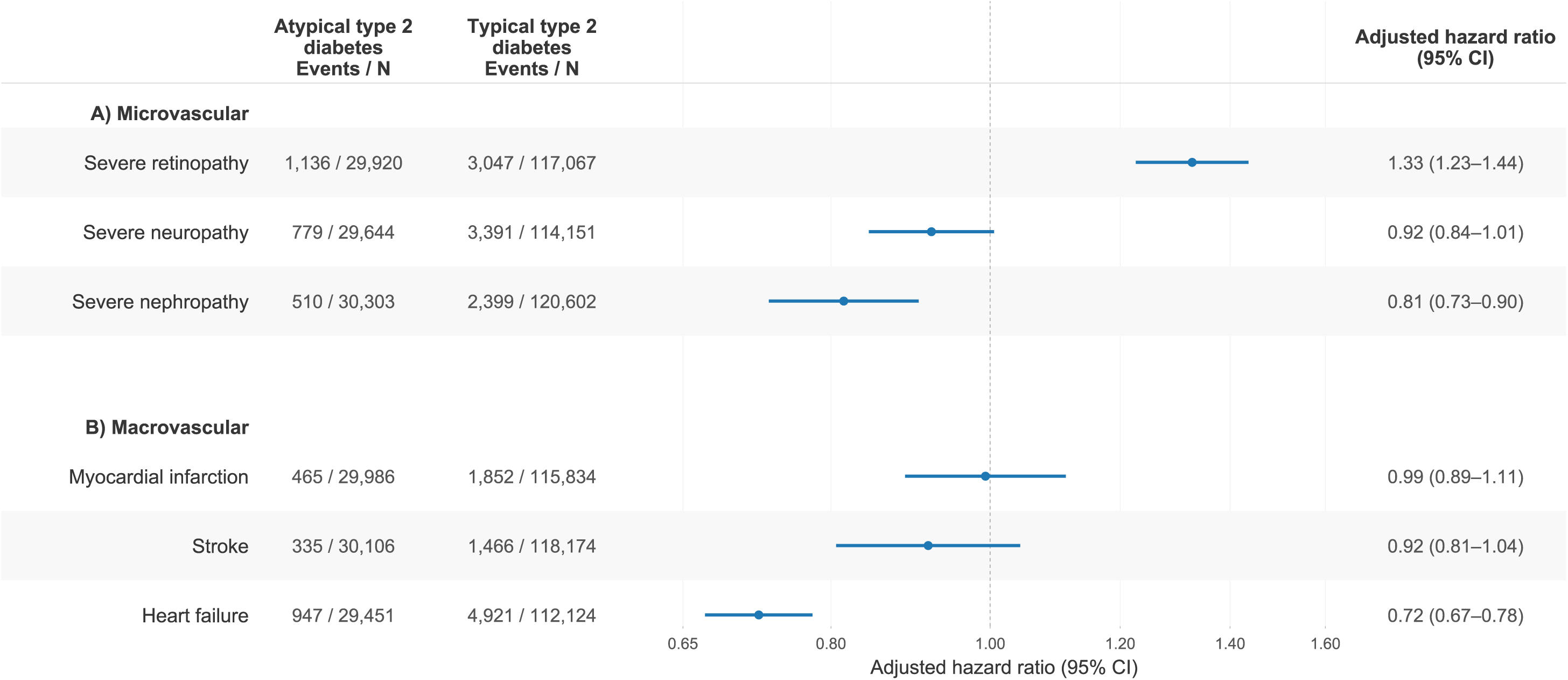
Adjusted hazard ratios for primary complications in atypical versus matched typical type 2 diabetes. Adjusted hazard ratios compare individuals with atypical type 2 diabetes (QDiabetes risk score ≤5.6%; n=30,392) with age-, sex- and calendar year-matched individuals with typical type 2 diabetes (QDiabetes risk score >5.6%; n=121,294). Outcomes were microvascular complications (severe retinopathy, neuropathy and nephropathy) and macrovascular complications (fatal or non-fatal myocardial infarction, stroke and heart failure). Models were adjusted for age at diagnosis, sex, calendar year of diagnosis, baseline HbA1c, ethnicity and deprivation. The matched typical type 2 diabetes group is the reference. Points indicate adjusted hazard ratios and horizontal lines indicate 95% confidence intervals; values >1 indicate higher risk in atypical type 2 diabetes.

There was no evidence of a difference in the risk of myocardial infarction (aHR 0.99, 95% CI 0.89 to 1.11) or stroke (aHR 0.92, 95% CI 0.81 to 1.04), while heart failure risk was lower in atypical type 2 diabetes (aHR 0.72, 95% CI 0.67 to 0.78). Findings for all complication outcomes were broadly consistent across age and sex subgroups (Supplementary Table 3).

### Atypical type 2 diabetes is associated with higher risks of acute glycaemic complications and non-cardiovascular mortality

After adjustment for sociodemographic factors and baseline HbA1c, atypical type 2 diabetes was associated with higher risks of diabetic ketoacidosis (aHR 1.66, 95% CI 1.29 to 2.15) and severe hypoglycaemia (aHR 1.24, 95% CI 1.05 to 1.46) (Table 2).

**Table 2.**
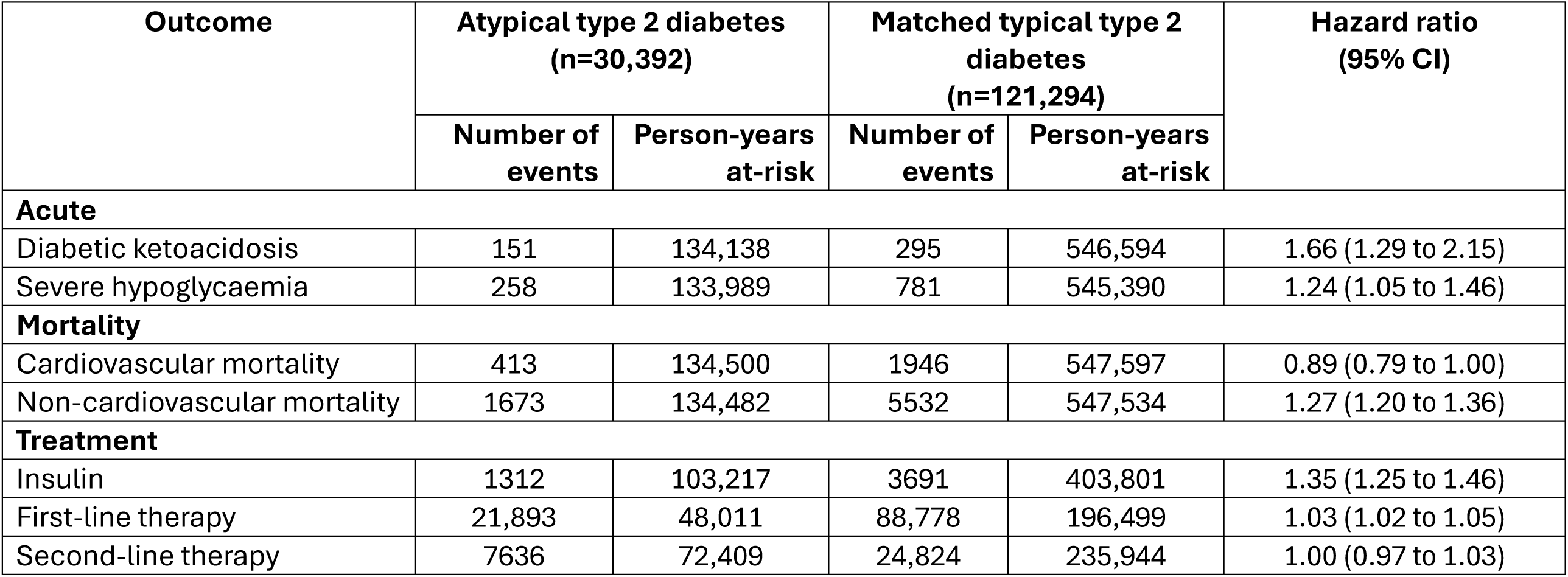
Secondary outcomes in atypical versus matched typical type 2 diabetes. . Event counts, person-years at risk and hazard ratios (95% CI) are shown for atypical type 2 diabetes (QDiabetes risk score ≤5.6%; n=30,392) and age-, sex- and calendar year-matched typical type 2 diabetes (QDiabetes risk score >5.6%; n=121,294). Models for acute complications and mortality were adjusted for age at diagnosis, sex, calendar year of diagnosis, baseline HbA1c, ethnicity and deprivation; treatment models were unadjusted. The matched typical type 2 diabetes group was the reference.

Atypical type 2 diabetes was associated with higher non-cardiovascular mortality (aHR 1.27, 95% CI 1.20 to 1.36), and there was no clear difference in risk of cardiovascular mortality (aHR 0.89, 95% CI 0.79 to 1.00) between groups. Underlying 10-year cumulative incidence plots are reported in Supplementary Figure 2.

### The atypical type 2 diabetes group progresses to insulin more quickly, although initiation of first- and second-line glucose-lowering therapy is similar

Insulin initiation in the 10-years after diagnosis was higher in atypical type 2 diabetes (11.3% [10.3 to 12.2] vs 8.9% [8.5 to 9.3]; HR 1.35, 95% CI 1.25 to 1.46) (Supplementary Figure 3; Table 2). In contrast, ten-year initiation of first-line glucose-lowering therapy was broadly similar between atypical and matched typical type 2 diabetes (87.5% [95% CI 86.6 to 88.4] vs 89.8% [89.4 to 90.2]; HR 1.03, 95% CI 1.02 to 1.05), as was second-line therapy initiation (64.6% [63.1 to 66.1] vs 67.6% [66.7 to 68.4]; HR 1.00, 95% CI 0.97 to 1.03).

## DISCUSSION

This large UK study demonstrates a clinically applicable approach to identify people with an atypical type 2 diabetes phenotype at diagnosis using routine clinical features. The group we identified was characterised by younger age, lower BMI, and higher initial HbA1c. Nevertheless, the phenotype was not restricted to younger or lean individuals, with cases also occurring in older adults and in people with obesity. Compared to age- and sex-matched typical type 2 diabetes counterparts, the atypical group had evidence of higher 10-year risks of retinopathy, diabetic ketoacidosis, severe hypoglycaemia, non-cardiovascular mortality and insulin initiation, that persisted after adjustment for HbA1c at diagnosis. Despite a more favourable baseline cardiovascular risk phenotype when assessed by conventional risk factors, the atypical group had similar 10-year risk of MI, stroke and cardiovascular mortality compared to individuals with typical type 2 diabetes.

Several factors may explain the clinical characteristics and adverse complication risk profile observed in the atypical type 2 diabetes group evaluated in this study. The combination of lower BMI, higher HbA1c at diagnosis, greater subsequent insulin requirement and increased retinopathy risk may be consistent with an underlying type 2 diabetes phenotype of greater beta-cell dysfunction,^8^ although specific biomarker measures would be required to directly assess this. The inadvertent inclusion of individuals with other types of diabetes, in particular misclassified type 1 or monogenic diabetes, could also contribute. However, our type 2 diabetes case definition excluded anyone who had ever had a type 1 diabetes clinical code or initiated insulin within one year of diagnosis, making substantial misclassification less likely, and meaning we may have in fact over-excluded atypical individuals with type 2 diabetes. Nonetheless, autoimmune or monogenic diabetes cannot be fully excluded without C-peptide, islet autoantibody and genetic testing.

For cardiovascular outcomes, the lower risk of heart failure was consistent with the more favourable baseline cardiometabolic profile of the atypical type 2 diabetes group, including lower BMI and less treated hypertension.^22 23^ In contrast, the similar risks of myocardial infarction and stroke were less expected given their lower baseline QRISK2 scores. Whilst our study was not designed to ascertain potential causes, a possible explanation is that the apparently more favourable risk profile of the atypical type 2 diabetes group may result in less intensive cardiovascular prevention. This was evident in our analysis where individuals with atypical type 2 diabetes had slightly higher LDL cholesterol, but a lower proportion were receiving statins at diagnosis. A related point is that the lower BMI and conventional cardiorenal risk of the atypical type 2 diabetes group may similarly make them less likely to be prioritised for newer glucose-lowering therapies with cardiovascular and renal benefits in current practice, including GLP-1 receptor agonists and SGLT2 inhibitors.^24 25^ Such inequalities and their impact could be assessed in future research.

Our findings correspond to a degree to recent diabetes subtyping studies that have proposed an insulin-deficient subgroup of type 2 diabetes.^7^ In particular, the atypical group evaluated here may overlap with the severe insulin-deficient diabetes (SIDD) subgroup identified in data-driven cluster analysis, which is similarly characterised by poor glycaemic control, earlier insulin requirement and increased retinopathy risk.^8 26 27^ Our analysis represents an advance on these cluster-based studies by firstly directly assessing complication outcomes against age- and sex-matched comparators, rather than across data-defined subgroups with very different age profiles, and secondly using routine features and not HOMA measures, which are not consistently available in clinical care.^6^

A further contrast can be made with young-onset type 2 diabetes (diagnosed <40 years of age) which is also associated with faster beta-cell decline and greater complication risk. However, young-onset diabetes is commonly associated with obesity, whereas individuals in the atypical group were leaner, and the phenotype was identified across all ages.^28^

This study has several strengths. We used a large, representative UK primary care cohort linked to hospital admissions and death records, allowing assessment of a broad range of severe clinical outcomes. Exact matching on age, sex and calendar year of diagnosis reduced major demographic and temporal confounding. However, as an observational study, we cannot determine causality and in particular it is possible that associations may reflect differences in clinical care including diagnostic delay, variation in healthcare testing, or management of diabetes post-diagnosis. However, differences in clinical outcomes in the atypical type 2 diabetes group persisted after adjustment for baseline HbA1c, and we found patterns of early initiation of glucose-lowering treatment post-diagnosis were broadly similar. As discussed above, diabetes misclassification may remain despite exclusion of coded type 1, type 3c and secondary diabetes. Finally, routine data lacked C-peptide and islet autoantibody measurements, so we could not directly assess beta-cell function, insulin secretion or autoimmune diabetes.

Importantly, and unlike clustering approaches whose limitations have been recently discussed,^29^ our analysis does not propose a discrete subtype of type 2 diabetes. Instead, our aim is to highlight the potential of repurposing existing widely used diabetes risk models to provide a simple way of identifying people with type 2 diabetes with a prognosis that differs from the conventional type 2 diabetes phenotype. The reproducibility and generalisability of this approach could be tested in other datasets, including with country-specific diabetes risk models, and utility could be tested in prospective studies to directly evaluate benefits of differential monitoring and treatment strategies.

Future research could also use the atypical group identified using our approach to inform genetic studies of underlying type 2 diabetes disease mechanisms. Partitioned polygenic risk scores capture distinct pathways, including beta-cell dysfunction, obesity, lipodystrophy and liver-lipid metabolism, and are associated with different clinical phenotypes and outcomes.^30 31^ Normal-weight individuals with type 2 diabetes have been shown to have higher beta-cell and lipodystrophy genetic risk but lower obesity-related genetic risk.^32^ Comparing pathway-specific scores between atypical and typical type 2 diabetes groups could assess to what degree beta-cell genetic risk contributes to the phenotype characterised by lower BMI and higher HbA1c observed in our study.

In conclusion, our study shows established type 2 diabetes onset risk prediction models identify people with atypical type 2 diabetes at diagnosis with a risk of complications that differs from typical type 2 diabetes. As such models are already widely used in routine practice worldwide, this approach could eventually provide a pragmatic approach to support more tailored management and treatment strategies for people with type 2 diabetes.

## Data Availability

Access to CPRD data is subject to protocol approval via CPRD's research data governance process (https://cprd.com/data-access). Code for initial cohort preparation is available at: https://github.com/Exeter-Diabetes/CPRD-Cohort-scripts/tree/main/01-At-diagnosis and analysis code is available at: https://github.com/Exeter-Diabetes/CPRD-Martha-atypical_T2D/tree/main

https://github.com/Exeter-Diabetes/CPRD-Martha-atypical_T2D/tree/main

## Contributors

The study concept and design were conceived and developed by MMD, JMD, BMS, TJM and KGY. MMD, JMD, KGY, PC, TTJ, and BMS had access to all the raw datasets used for the study. APM reviewed the clinical codes. MMD undertook the analysis, with support from KGY, TJM, BMS and JMD. All authors provided support for the analysis and interpretation of results, critically revised the manuscript, and approved the final manuscript. MMD and JMD attest that all listed authors meet authorship criteria and that no others meeting the criteria have been omitted. All authors had final responsibility for the decision to submit for publication.

## Transparency

The lead author (JMD) is the guarantor of this manuscript and affirms that the manuscript is an honest, accurate, and transparent account of the study being reported; that no important aspects of the study have been omitted; and that any discrepancies from the study as originally planned have been explained. The study protocol was not pre-registered.

## Funding

This research was supported by the Wellcome Trust (227070/Z/23/Z), Medical Research Council (UK) (MR/W003988/1). This study has been delivered through the National Institute for Health and Care Research (NIHR) Exeter Biomedical Research Centre (BRC) (NIHR203320). The funders had no role in considering the study design or in the collection, analysis, interpretation of data, writing of the report, or decision to submit the article for publication. The views expressed are those of the author(s) and not necessarily those of the Wellcome Trust, Medical Research Council, the NIHR or the Department of Health and Social Care. For the purpose of open access, the authors have applied a Creative Commons Attribution (CC BY) licence to any Author Accepted Manuscript version arising from this submission.

## Declaration of interests

JMD is supported by a Wellcome Trust Early Career award (227070/Z/23/Z). ATH and BMS are supported by the NIHR Exeter Clinical Research Facility; the views expressed are those of the authors and not necessarily those of the NHS, the NIHR or the Department of Health. APM declares previous research funding from Eli Lilly, Pfizer, and AstraZeneca. AGJ declares research funding to his university from the UK Medical Research Council, NIHR, Diabetes UK, Breakthrough Type 1 diabetes, the Novo Nordisk Foundation and European Foundation for the Study of Diabetes. LMG is supported by the Engineering and Physical Sciences Research Council (EPSRC) through the Causality in Healthcare AI Hub (CHAI) (EP/Y028856/1).

## Ethical approval

This study was approved by the CPRD independent scientific advisory committee (eRAP 24_004747). CPRD also has ethical approval from the Health Research Authority to support research using anonymised patient data (research ethics committee reference 21/EM/0265). Individual patient consent was not required as all data were deidentified.

## Notes

### Competing Interest Statement

The authors have declared no competing interest.

