## Supplementary Table 1 for "A model-based approach to identifying atypical type 2 diabetes: UK population-based study"

**Supplementary Flowchart**. Derivation of the study cohort from adults newly diagnosed with type 2 diabetes in CPRD Aurum, 2013 to 2023.

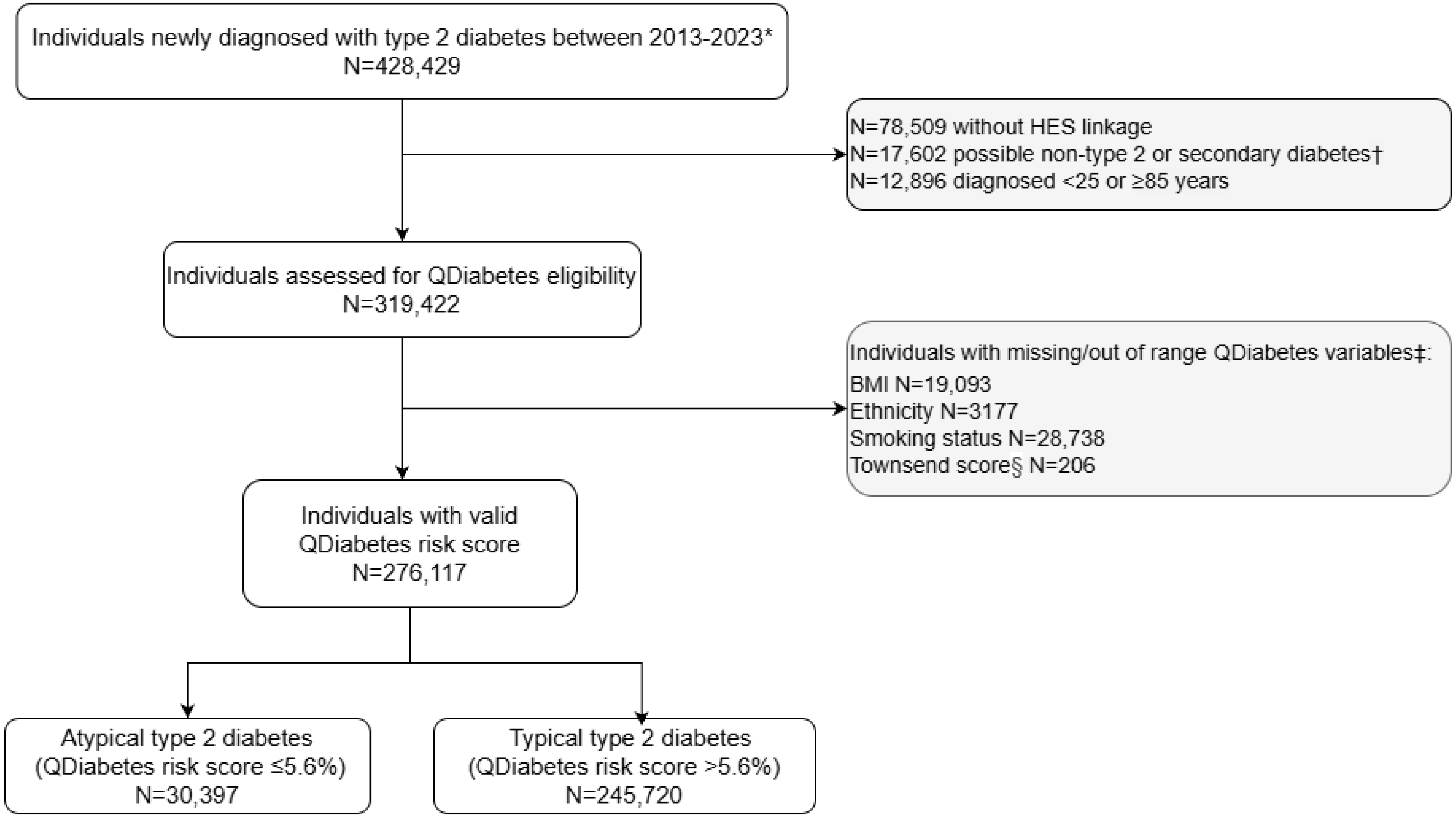


*Including only individuals diagnosed at least 90 days after practice registration.

†Alternative or secondary causes of diabetes were defined as evidence of type 1 diabetes (insulin initiation within 1 year of diagnosis or any recorded type 1 diabetes code), type 3c diabetes (history of acute/chronic pancreatitis, pancreatic cancer, pancreatic resection, or haemochromatosis), or cystic fibrosis.

‡All other QDiabetes predictors had complete data. Individuals could contribute to more than one missing/out-of-range QDiabetes variable.

§Townsend deprivation score (TDS) missing or out of bounds (<−8 or >14). To estimate TDS scores from IMD, we used 2019 IMD deciles and 2011 TDS scores for all Lower Super Output Areas (LSOAs) in England and found the median 2011 TDS corresponding to each IMD decile.

HES = Hospital Episode Statistics

**Supplementary Table 1**. **Baseline characteristics in atypical versus unmatched typical type 2 diabetes.** Individuals with atypical type 2 diabetes (QDiabetes risk score ≤5.6%; n=30,397) were compared with individuals with typical type 2 diabetes (QDiabetes risk score >5.6%; n=245,720). Values are n (%) for categorical variables and mean [SD] for continuous variables. Percentages for variables with missing data are calculated among individuals with available data. Standardised mean differences (SMDs) compare the atypical type 2 diabetes group with the unmatched typical type 2 diabetes group, SMD >0.1 indicates meaningful imbalance.

| | | **Atypical type 2 diabetes (n=30,397)** | **Unmatched typical type 2 diabetes (n=245,720)** | **SMD** |
|---|---|---|---|---|
| | Sex (% male) | 14,871 (48.9) | 145,670 (59.3) | 0.21 |
| | Age at diagnosis (years) | 53.8 [15.9] | 59.9 [12.1] | 0.43 |
| **Age at diagnosis category (%)** | 25-39 | 6785 (22.3) | 12,320 (5.0) | 0.63 |
| | 40-49 | 6932 (22.8) | 41,194 (16.8) | |
| | 50-59 | 6110 (20.1) | 70,906 (28.9) | |
| | 60-69 | 4432 (14.6) | 65,140 (26.5) | |
| | 70-79 | 4163 (13.7) | 44,822 (18.2) | |
| | 80-84 | 1975 (6.5) | 11,338 (4.6) | |
| **Ethnicity (%)** | White | 24,277 (79.9) | 193,423 (78.7) | 0.14 |
| | South Asian | 2874 (9.5) | 31,655 (12.9) | |
| | Black | 1852 (6.1) | 13,602 (5.5) | |
| | Mixed | 488 (1.6) | 2664 (1.1) | |
| | Other | 906 (3.0) | 4376 (1.8) | |
| **IMD quintile (%)** | 1 (least deprived) | 6577 (21.6) | 37,589 (15.3) | 0.24 |
| | 2 | 6309 (20.8) | 43,176 (17.6) | |
| | 3 | 5964 (19.6) | 46,863 (19.1) | |
| | 4 | 6085 (20.0) | 56,294 (22.9) | |
| | 5 (most deprived) | 5462 (18.0) | 61,798 (25.1) | |
| **Smoking category (%)** | Active smoker | 6345 (20.9) | 57,037 (23.2) | 0.26 |
| | Ex-smoker | 12,498 (41.1) | 124,616 (50.7) | |

| | | | | |
|---|---|---|---|---|
| | Non-smoker | 11,554 (38.0) | 64,067 (26.1) | |
| **Alcohol category (%)** | Excess | 2628 (10.4) | 23,762 (10.9) | 0.04 |
| | Harmful | 1081 (4.3) | 11,058 (5.1) | |
| | Within limits | 18,770 (74.2) | 159,490 (73.1) | |
| | None | 2810 (11.1) | 23,865 (10.9) | |
| **Clinical features and laboratory measurements** | Family history of diabetes | 4728 (15.6) | 37,097 (15.1) | 0.01 |
| | eGFR (mL/min/1.73 $m^2$) | 95 [19.7] | 88 [18.5] | 0.38 |
| | HDL-c(mmol/L) | 1.3 [0.4] | 1.2 [0.3] | 0.28 |
| | LDL-c (mmol/L) | 3.2 [1.0] | 2.9 [1.0] | 0.30 |
| | TG/HDL-c ratio | 2.3 [3.9] | 2.3 [2.7] | <0.01 |
| | QDiabetes 10-year risk score | 3.7 (1.3) | 21.6 (14.0) | 1.8 |
| | QRISK2 10-year risk score* | 16.7 [14.8] | 25.4 [14.3] | 0.60 |
| **HbA1c (mmol/mol) †** | Mean (SD) | 68.0 [24.9] | 60.0 [18.5] | 0.36 |
| **HbA1c category, mmol/mol (%)** | 48-58 | 15,400 (55.9) | 157,202 (69.6) | 0.36 |
| | 59-74 | 3952 (14.4) | 33,903 (15.0) | |
| | 75-85 | 1745 (6.3) | 9725 (4.3) | |
| | ≥86 | 6442 (23.4) | 24,946 (11.0) | |
| **BMI (kg/$m^2$)** | Mean (SD) | 26.4 [5.1] | 34.0 [6.9] | 1.25 |
| **BMI category, kg/$m^2$ (%)** | <25 | 12,870 (42.3) | 10,514 (4.3) | 1.45 |
| | 25-29 | 12,731 (41.9) | 64,042 (26.1) | |
| | 30-39 | 4131 (13.6) | 130,940 (53.3) | |
| | ≥40 | 665 (2.2) | 40,224 (16.4) | |
| **Comorbidities (%)** | Cardiovascular disease‡ | 2262 (7.4) | 54,524 (22.2) | 0.42 |
| | Myocardial infarction | 406 (1.3) | 12523 (5.1) | 0.22 |
| | Stroke | 286 (0.9) | 5166 (2.1) | 0.09 |
| | Heart failure | 941 (3.1) | 14,528 (5.9) | 0.14 |
| | Hypertension requiring treatment | 3359 (11.1) | 111,858 (45.5) | 0.83 |

| | | | | |
|---|---|---|---|---|
| | Chronic kidney disease stages 3-5 | 1525 (5.0) | 20,306 (8.3) | 0.13 |
| | Polycystic ovary syndrome | 345 (1.1) | 2498 (1.0) | 0.01 |
| | Bipolar disorder or schizophrenia | 350 (1.2) | 7079 (2.9) | 0.12 |
| | Learning disabilities | 430 (1.4) | 2566 (1.0) | 0.03 |
| **Treatment history (%)** | Oral steroids | 1768 (5.8) | 26,528 (10.8) | 0.18 |
| | Statins | 3334 (11.0) | 96,055 (39.1) | 0.69 |
| | Antipsychotic medication | 748 (2.5) | 14,066 (5.7) | 0.17 |

* QRISK2 was calculated among individuals without pre-existing cardiovascular disease and complete data for all required variables (n=216,290)

† HbA1c was missing for 22,802 (8.3%) individuals

‡Cardiovascular disease: myocardial infarction, stroke, revascularisation, ischaemic heart disease, angina, peripheral arterial disease, transient ischaemic attack

**Supplementary Table 2. Primary outcomes in atypical versus matched typical type 2 diabetes.** Event counts, person-years at risk and adjusted hazard ratios (95% CI) are shown for atypical type 2 diabetes (QDiabetes risk score ≤5.6%; n=30,392) and age-, sex- and calendar year-matched typical type 2 diabetes (QDiabetes risk score >5.6%; n=121,294). Models were adjusted for age at diagnosis, sex, calendar year of diagnosis, baseline HbA1c, ethnicity and deprivation. The matched typical type 2 diabetes group was the reference.

| Outcome | Atypical type 2 diabetes (n=30,392) | | Matched typical type 2 diabetes (n=121,294) | | Adjusted hazard ratio (95% CI) |
|---|---|---|---|---|---|
| | Number of events | Person-years at-risk | Number of events | Person-years at-risk | |
| **Microvascular** | | | | | |
| Severe retinopathy | 1136 | 129,217 | 3047 | 520599 | 1.33 (1.23 to 1.44) |
| Severe neuropathy | 779 | 129003 | 3391 | 506288 | 0.92 (0.84 to 1.01) |
| Severe nephropathy | 510 | 133004 | 2399 | 539760 | 0.81 (0.73 to 0.90) |
| **Macrovascular** | | | | | |
| Myocardial infarction | 465 | 131731 | 1852 | 519487 | 0.99 (0.89 to 1.11) |
| Stroke | 335 | 132715 | 1466 | 531496 | 0.92 (0.81 to 1.04) |
| Heart failure | 947 | 129302 | 4921 | 498860 | 0.72 (0.67 to 0.78) |

**Supplementary Table 3. Adjusted hazard ratios for primary complications in atypical versus matched typical type 2 diabetes, overall and stratified by age and sex.** Adjusted hazard ratios compare individuals with atypical type 2 diabetes (QDiabetes risk score ≤5.6%) with age-, sex- and calendar year-matched individuals with typical type 2 diabetes (QDiabetes risk score >5.6%). Outcomes were microvascular complications (severe retinopathy, neuropathy and nephropathy) and macrovascular complications (fatal or non-fatal myocardial infarction, stroke and heart failure). Results are shown overall and stratified by age at diagnosis (<50 or ≥50 years) and sex. Models were adjusted for age at diagnosis, sex, calendar year of diagnosis, ethnicity, deprivation and baseline HbA1c. The matched typical type 2 diabetes group is the reference.

| **Outcome** | **Overall study population aHR (95% CI)** | **Age at diagnosis <50 years aHR (95% CI)** | **Age at diagnosis ≥50 years aHR (95% CI)** | **Female aHR (95% CI)** | **Male aHR (95% CI)** |
|---|---|---|---|---|---|
| Atypical type 2 diabetes, n | 30,392 | 13,712 | 16,680 | 15,521 | 14,871 |
| Matched typical type 2 diabetes, n | 121,294 | 54,704 | 66,590 | 61,902 | 59,392 |
| **Microvascular** | | | | | |
| Severe retinopathy | 1.33 (1.23 to 1.44) | 1.35 (1.16 to 1.57) | 1.31 (1.20 to 1.44) | 1.25 (1.12 to 1.39) | 1.43 (1.28 to 1.60) |
| Severe neuropathy | 0.92 (0.84 to 1.01) | 0.92 (0.78 to 1.09) | 0.91 (0.82 to 1.01) | 0.88 (0.78 to 0.99) | 0.96 (0.85 to 1.09) |
| Severe nephropathy | 0.81 (0.73 to 0.90) | 0.71 (0.52 to 0.95) | 0.82 (0.73 to 0.92) | 0.74 (0.65 to 0.86) | 0.94 (0.80 to 1.09) |
| **Macrovascular** | | | | | |
| Myocardial infarction | 0.99 (0.89 to 1.11) | 0.83 (0.65 to 1.06) | 1.05 (0.93 to 1.19) | 0.98 (0.82 to 1.16) | 1.00 (0.86 to 1.16) |
| Stroke | 0.92 (0.81 to 1.04) | 0.66 (0.45 to 0.98) | 0.97 (0.85 to 1.11) | 0.89 (0.74 to 1.06) | 0.96 (0.79 to 1.15) |
| Heart failure | 0.72 (0.67 to 0.78) | 0.71 (0.57 to 0.89) | 0.72 (0.67 to 0.78) | 0.73 (0.66 to 0.81) | 0.72 (0.64 to 0.80) |

**Supplementary Figure 1. Clinical characteristics at diagnosis in atypical versus unmatched typical type 2 diabetes**. Distributions of age, body mass index (BMI), HbA1c and triglyceride-to-HDL cholesterol (TG/HDL) ratio are shown for individuals with atypical type 2 diabetes (QDiabetes risk score ≤5.6%; n=30,397) and unmatched typical type 2 diabetes (QDiabetes risk score >5.6%; n=245,720). Violin plots show the distribution of each characteristic; box plots show the median and interquartile range.

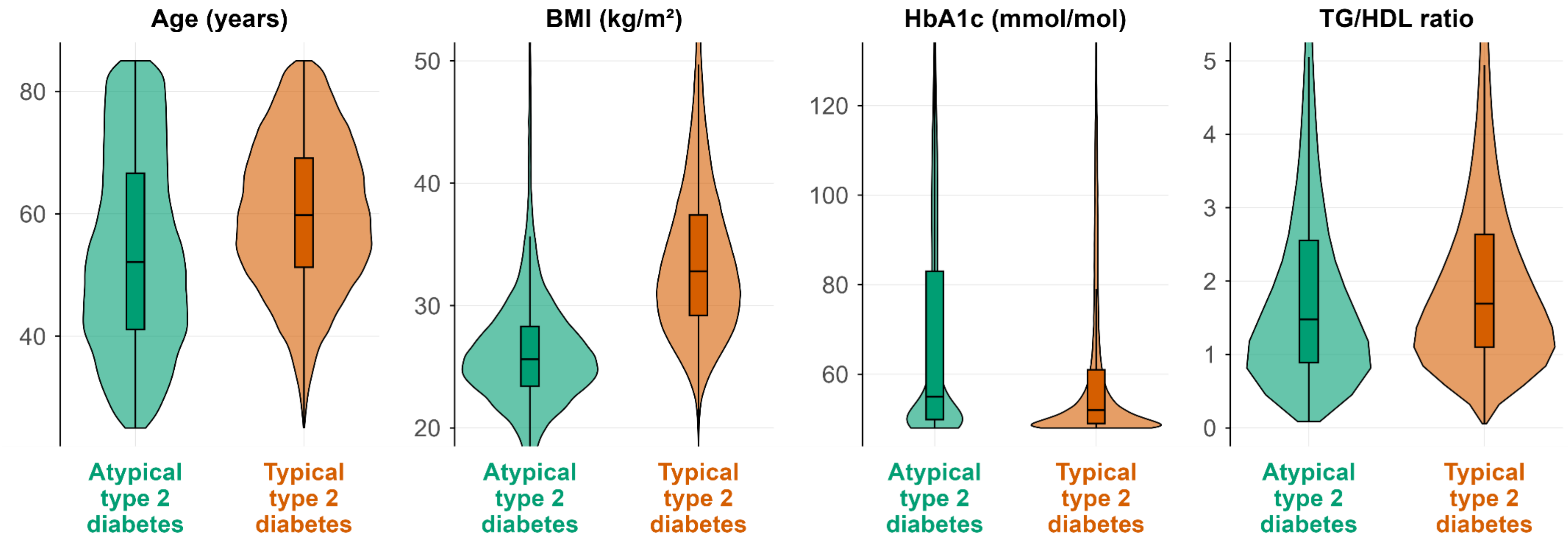

**Supplementary Figure 2. Ten-year cumulative incidence of secondary outcomes in atypical versus matched typical type 2 diabetes.** Kaplan–Meier curves compare individuals with atypical type 2 diabetes (QDiabetes risk score ≤5.6%; n=30,392) with age-, sex- and calendar year-matched individuals with typical type 2 diabetes (QDiabetes risk score >5.6%; n=121,294) for acute complications (diabetic ketoacidosis and severe hypoglycaemia) and mortality (cardiovascular and non-cardiovascular). Shaded areas indicate 95% confidence intervals; numbers below each panel indicate participants at risk.

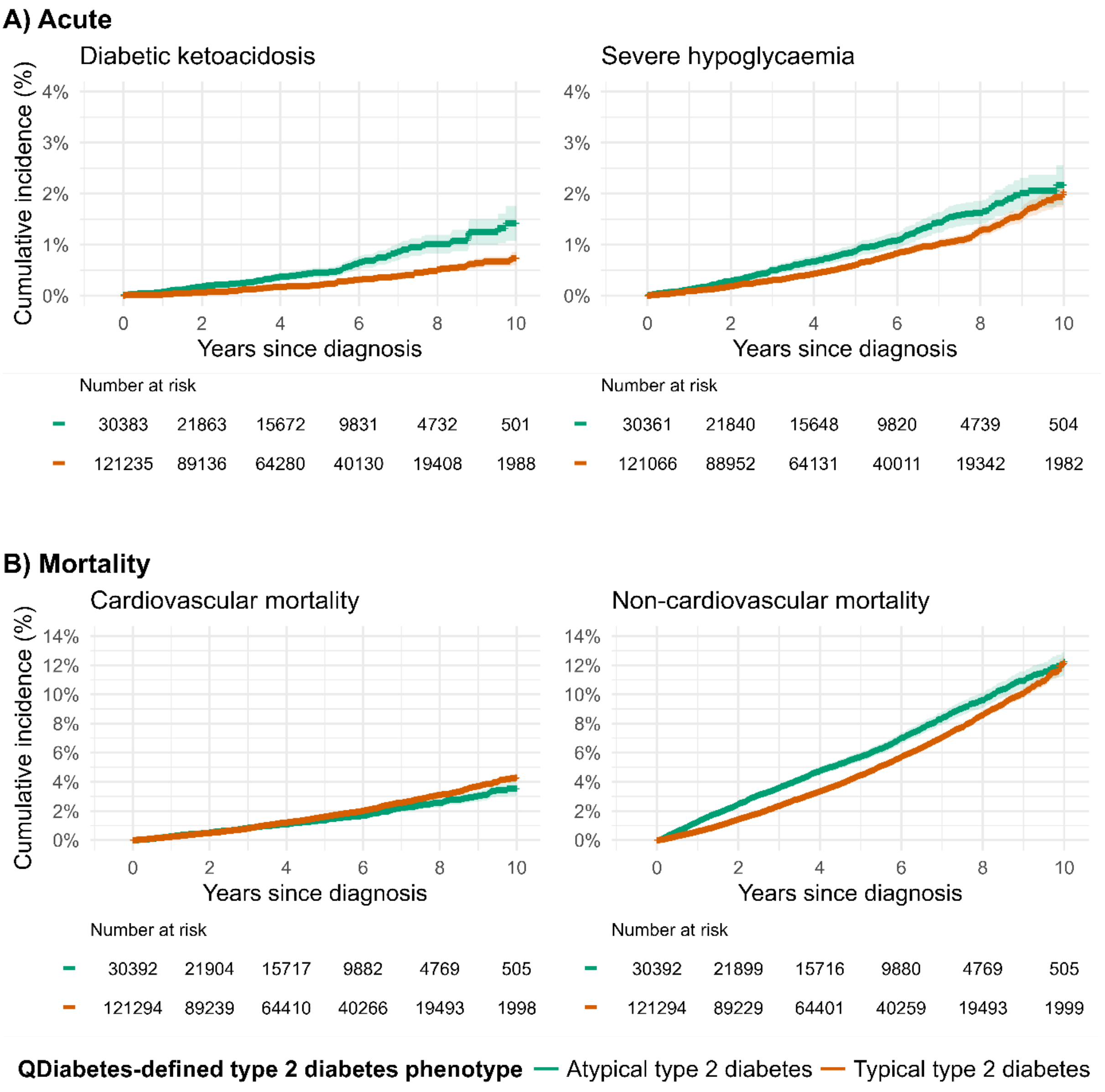

**Supplementary Figure 3. Ten-year cumulative incidence of glucose-lowering treatment initiation in atypical versus matched typical type 2 diabetes.** Kaplan–Meier curves compare individuals with atypical type 2 diabetes (QDiabetes risk score ≤5.6%; n=30,392) with age-, sex- and calendar year-matched individuals with typical type 2 diabetes (QDiabetes risk score >5.6%; n=121,294) for insulin initiation, first-line glucose-lowering therapy initiation, and second-line therapy added to initial metformin. Follow-up for insulin initiation began 1 year after diagnosis. Shaded areas indicate 95% confidence intervals; numbers below each panel indicate participants at risk.

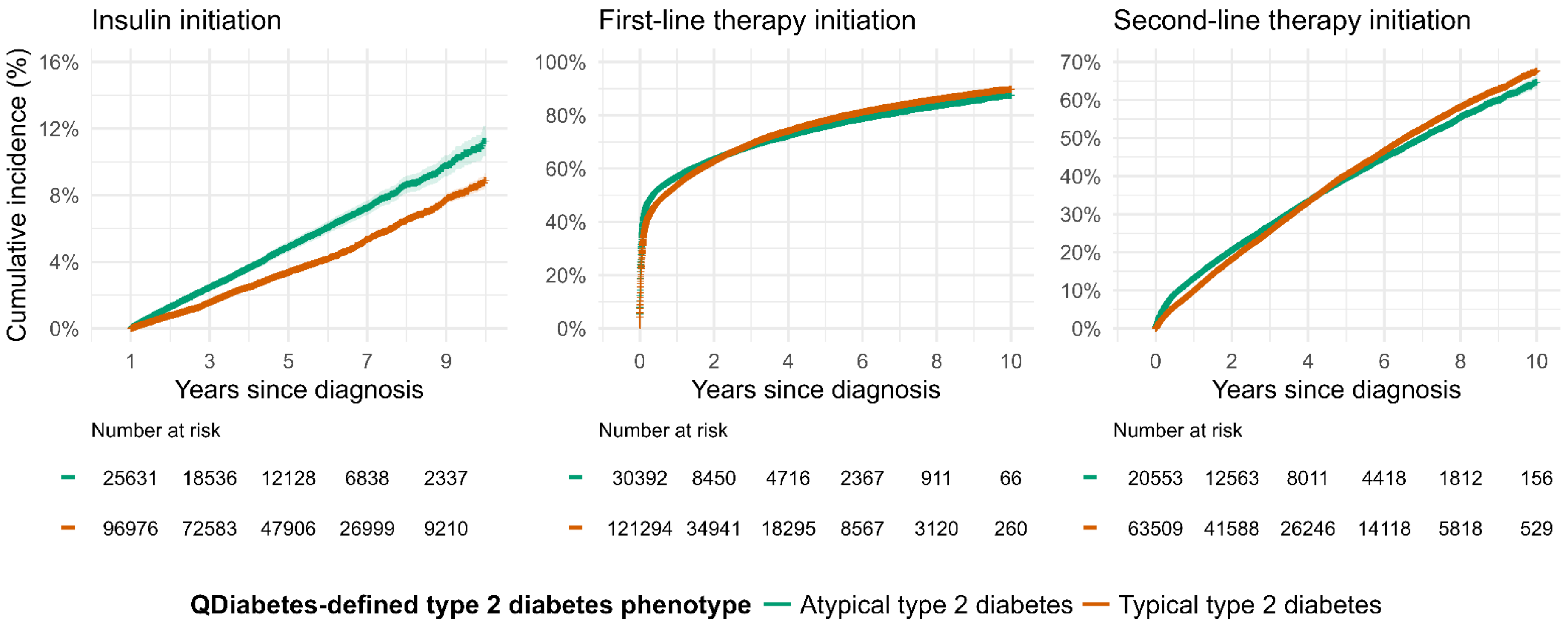
